# The XForce Tourniquet: A Comparative Analysis with the CAT Tourniquet to Advance Efficacy and Establish Foundations for Smart Hemorrhage Control

**DOI:** 10.1101/2025.03.15.25324011

**Authors:** Altobelli Anthony, Pai Esha, Bandaru Aishwarya, Yanamala Naveena

## Abstract

**Background:** Tourniquets have demonstrated life-saving efficacy within military settings as essential tools in hemorrhage control. Despite their proven effectiveness, traditional windlass-based tourniquets such as the Combat Application Tourniquet (CAT) present challenges in rapid application and ease of use, particularly within civilian emergency contexts. The XForce Tourniquet (XForce TQ) has been developed to address these limitations with a novel ratcheting mechanism and self-securing strap. These design features aim to improve usability and application speed while also demonstrating the XForce tourniquets’ ability to serve as the foundation for broader telemedicine tourniquet initiatives.

**Methods:** This study conducted a comparative evaluation of the XForce TQ and CAT TQ among healthcare professionals (n = 99) using a simulated limb model (TQ Aid). Participants applied both tourniquets in three timed trials each with application times recorded at key steps. The study assessed differences in mean total application time, user performance across age and sex groups, and overall device efficiency. Statistical analyses included paired t-tests and ANOVA to determine significance.

**Results:** The XForce TQ significantly reduced mean total application time (8.67 ± 2.12 s) compared to the CAT TQ (16.53 ± 4.43 s, p < 0.001), representing a 47% reduction in total application time. Significant differences were also observed between sexes, with females taking longer to apply both tourniquets (p < 0.05). No significant differences in application time were found between age groups (p = 0.852). The ratcheting mechanism of the XForce TQ demonstrated improved user efficiency and reduced application variability.

**Conclusion:** The XForce TQ offers significantly faster application times than the CAT TQ, suggesting that its novel design enhances usability in emergency scenarios. These findings support the development of next-generation intelligent tourniquets integrating smart features such as automated emergency alerts and telemedicine capabilities. Further research is needed to validate its performance in real-world trauma settings.

## Introduction

The use of tourniquets as a critical care intervention for hemorrhage control has a well-documented history, particularly within military medicine. The conflicts in Iraq and Afghanistan throughout the early 2000s provided pivotal evidence demonstrating the life-saving potential of tourniquets in managing catastrophic extremity hemorrhage. Extensive data from these conflicts revealed that the reintroduction of tourniquets on the battlefield led to significant reductions in mortality rates among military personnel. Notably, studies have demonstrated that prehospital application of tourniquets was instrumental in preventing exsanguination when applied promptly, thereby reducing fatalities from extremity injuries.^1^ The Tactical Combat Casualty Care (TCCC) guidelines, which were refined and rigorously implemented during these conflicts, prioritized the use of tourniquets as a first-line intervention. Analyses of combat casualty data revealed that adherence to these protocols produced an association with a marked reduction in preventable deaths due to limb hemorrhage.^2^ Furthermore, research conducted during Operation Iraqi Freedom demonstrated that early application of tourniquets improved outcomes in severely injured patients, with no significant increase in the advent of limb ischemia or need for secondary amputation.^3^

The demonstrated efficacy of tourniquets in military settings has led to increased interest in their adoption within civilian trauma care, particularly in response to the increasing frequency of mass-casualty incidents, such as active shooter events. Recognizing the need for immediate hemorrhage control in civilian settings, the “Stop the Bleed” Campaign was launched in 2015 by The White House and supported by the American College of Surgeons (ACS). The initiative aimed to train civilians to act as immediate responders to severe bleeding incidents, thereby bridging the critical time gap before professional medical help arrives.^4^ The campaign has successfully trained over 125,000 individuals in hemorrhage control techniques, significantly enhancing public preparedness in the advent of traumatic bleeding.^5^ Such public training initiatives have been shown to improve bystanders’ ability to control bleeding and potentially save patient lives during emergency medical situations.^6^

In response to these developments, various commercially available tourniquet devices have been employed in civilian prehospital settings to enhance trauma care. Among these, the Combat Application Tourniquet (CAT) has become the standard due to its provided efficacy in controlling hemorrhage.^7^ Studies have demonstrated that the CAT is effective in both military and civilian settings, with effectiveness rates ranging from 78 to 100%.^7^ During the Boston Marathon bombing, the CAT tourniquet’s reliability was highlighted when applied by trained professionals and civilians alike.^8^ However, while the CAT has demonstrated effectiveness in preventing hemorrhage when applied correctly, inconsistencies in its application in civilian settings remain a challenge. These inconsistencies often arise due to inadequate training, multi-step application complexity, and potential design limitations. Studies indicate that even trained responders may struggle with the correct application of the CAT due to its design, which can result in improper tightening and reduced effectiveness.^9^ Research has also demonstrated that modifications to the CAT, such as a slack-reducing band, can improve its pressure application and hemorrhage control rates, suggesting the current mechanical drawbacks of current CAT models.^10^ Furthermore, studies have highlighted that the CAT’s bulkiness and complexity can hinder rapid application, especially in high-stress civilian circumstances where responders may not possess the same level of training as military personnel.^11^ These findings underscore the need for improved training protocols and potential design refinements to optimize the tourniquet’s performance in diverse, non-military environments. While the CAT tourniquet is widely used, challenges related to application consistency, speed, and user training highlight the need for more intuitive and efficient designs. This study aims to evaluate the XForce Tourniquet as a potential solution to these limitations.

Thus, this study explores the potential of a next-generation intelligent tourniquet system to enhance trauma care by improving application speed and usability while incorporating advanced smart features. The XForce tourniquet, designed to overcome key limitations of the widely used CAT tourniquet, features an intuitive ratcheting mechanism and a self-securing strap, reducing the complexity of application. By comparing the XForce tourniquet’s efficiency to the CAT, this study aims to establish a foundation for integrating smart hemorrhage control technologies into trauma care.

To further assess its suitability as a platform for next-generation smart tourniquet systems, we conducted a comparative evaluation of the XForce and CAT tourniquets among healthcare providers. Specifically, this study examines application times and user experience to determine whether the XForce design enhances usability and effectiveness. These findings will support future integration of real-time GPS tracking, automated emergency alerts, and telemedicine capabilities into the TeleTQ system—a next-generation smart tourniquet aimed at improving both hemorrhage control and emergency response coordination.

## Methods

### Xforce Tourniquet (XForce TQ)

The Xforce TQ (Figure 1), developed by Auric Innovations, is tested within this study to determine the capability of the Xforce TQ to act as the foundational component for a future Xforce TELE-TQ system. The Xforce incorporates a series of design advancements aimed at improving ease of use, application speed, and overall effectiveness in hemorrhage control. Unlike traditional windlass–based tourniquets, the Xforce employs a ratcheting lever mechanism (Figure 1a). The ratcheting mechanism provides a mechanical advantage by allowing users to apply controlled pressure with minimal effort and dexterity. This is meant to simplify the tightening process with the hope of ensuring consistent pressure application throughout the hemorrhagic crisis. The lever complex also features dual release levers (Figure 1a). The dual release levers are a proprietary, patented technology that allows for gradual pressure release to safely transition the tourniquet into a pressure dressing. Ability to safely and effectively convert from tourniquet use to application of a pressure dressing is associated with prevention of rebleeding at the injury site while promoting healing. The XForce TQ features a zip-tie-inspired strap for simplified application. The design allows for users to simply pull the strap to tighten. With the strap, it automatically locks in place which eliminates additional steps and reduces the risk of user error when securing the tourniquet. Unlike traditional windlass-based tourniquets, the XForce TQ’s ratcheting lever mechanism requires less dexterity and effort, making it more accessible for users with limited training or high-stress conditions.

**Figure 1.**
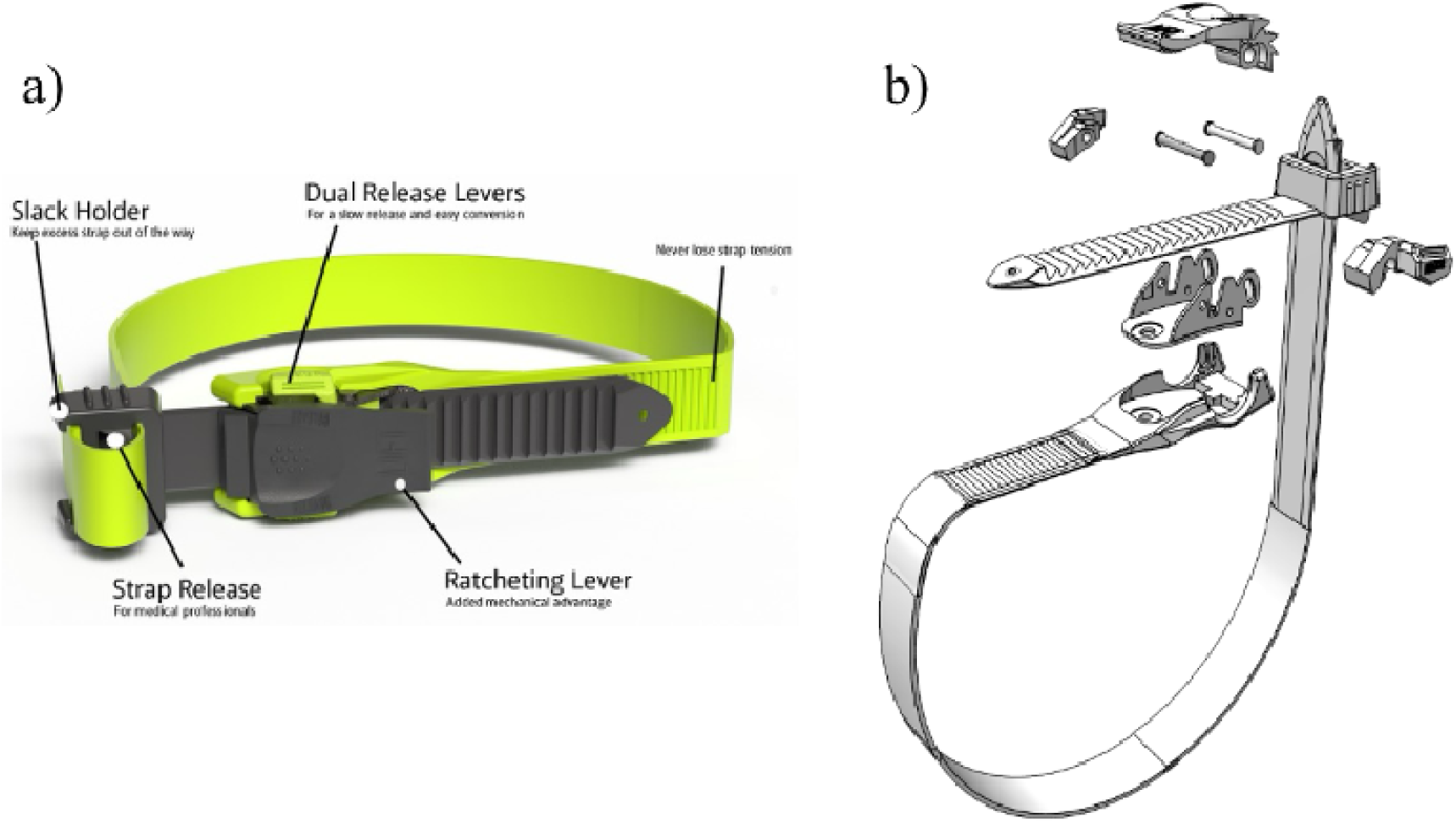
a) XForce tourniquet unique design implementations and b) a blown up schematic to highlight the design and integration details of various components.

The Xforce design features efficiency in hemorrhagic control as well as production. Over 85% of its components are produced using plastic injection molding, ensuring consistency, rapid production, and cost-effectiveness, especially considering its production within the USA. The benefits of injection molding provide the capacity for high-volume production with uniformity in regard to the quality. This combination provides the capability for scalable manufacturing at a reduced cost. Additionally, the Xforce TQ is engineered for quick and simple assembly with each unit interlocking efficiently. This streamlined process enhances efficiency, reliability, and accessibility for users within emergency situations.

### TQ AID Simulator

To ensure adequate pressure was achieved, during participant applications of both the CAT and Xforce TQs, the TQ Aid (Figure 2a), created by Sim Limb, was employed. The TQ Aid serves as a baseline simulator for effective tourniquet application. Constructed from proprietary silicone, it mimics the feel of a human limb and contains a pressure sensor calibrated to indicate complete arterial occlusion when applied pressure exceeds 500 mmHg. Before adequate pressure is achieved, the TQ Aid features an LED light that when displaying red indicates inadequate applied pressure (P_Applied_<500 mmHg) (Figure 2b). Once P_Applied_ >500 mmHg the LED light turns green (Figure 2c). Progression of applied pressure during application can also be monitored on the LED screen found on the right side of the limb. For the purpose of this non-human, comparative study, the pressure required for the TQ Aid pressure sensor to indicate complete arterial occlusion is not meant to reflect clinically recommended pressures. Rather, the 500 mmHg serves as an arbitrary marked point purposefully chosen to be higher than the desired pressure for clinical contexts.

**Figure 2.**
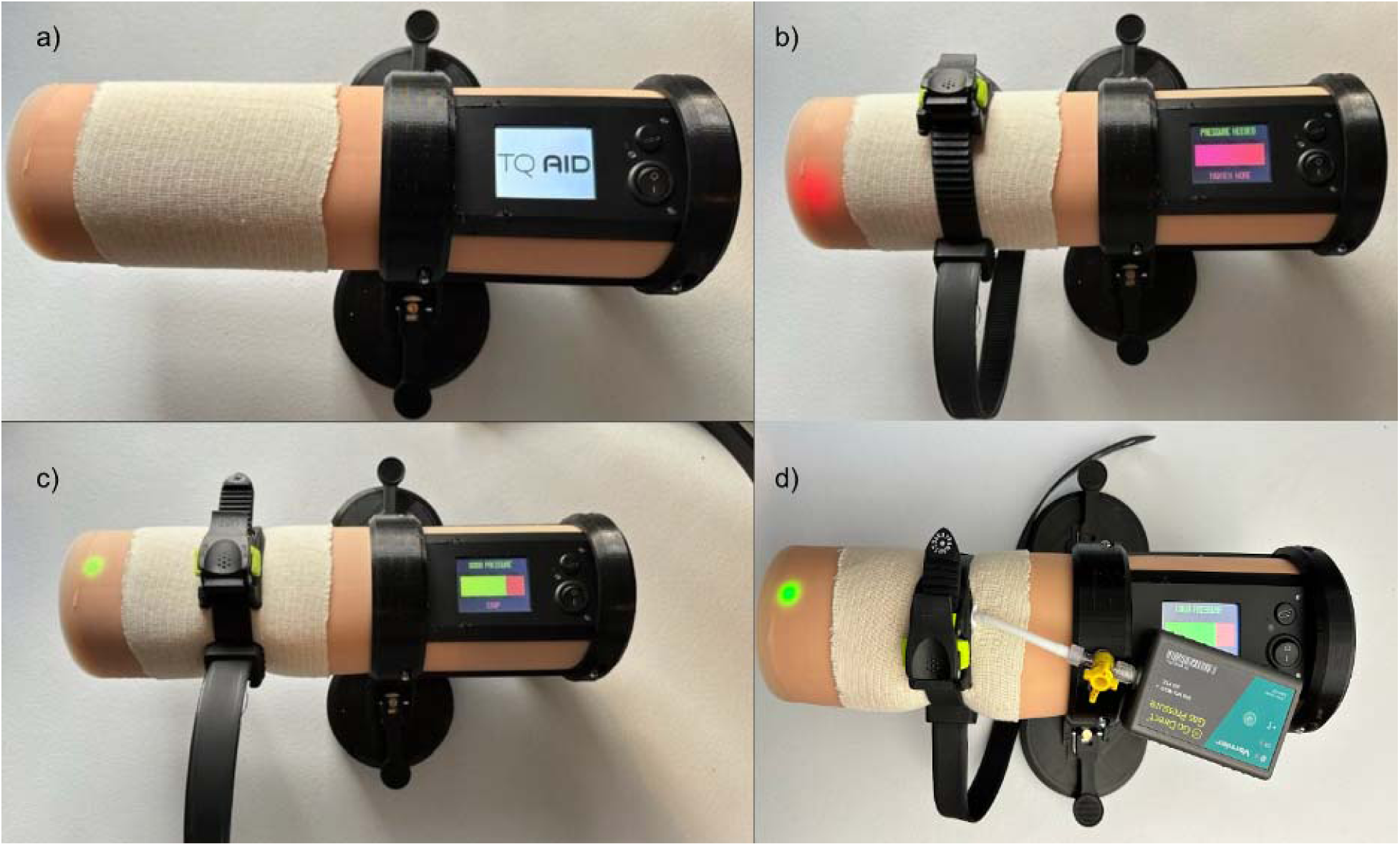
a) The TQ Aid developed by sim limb used to monitor application pressure during participant application. b) XForce tourniquet resting on TQ Aid demonstrating inadequate pressure. c) XForce tourniquet fully applied on TQ Aid demonstrating adequate pressure. d) The Vernier Go Direct Gas Pressure Sensor affixed to TQ Aid under XForce TQ. The CAT TQ employed the same set up to monitor application pressure.

While designed to indicate adequate applied pressure at 500 mmHg, further confirmation was achieved through a series of limb calibration tests. The Go Direct Gas Pressure Sensor by Vernier was affixed to the TQ Aid allowing for determination of the average pressure required for the TQ Aid to light green (Figure 2d). A total of n=20 applications were conducted with both the XForce TQ and CAT TQ contributing 10 applications each. The data collection was recorded through a vernier software which displayed applied pressure in mmHg as a function of time. The time to reach maximum applied pressure was recorded as well as the time necessary to reach said pressure.

## Comparative Analysis of CAT and Xforce TQ Application

### Study Participation

To evaluate the feasibility of the XForce TQ prototype as a viable candidate for future intelligent tourniquet technology, a non-human, comparative testing phase was conducted. A total of n=99 participants were enrolled into the study which exceeds the recommended n=75 requirement for statistical significance to be achieved. Healthcare professionals of all ranks were invited to participate in the study on a free-will basis. Once a participant indicated a desire to participate, they were briefed on the purpose of the study and its entailed procedures before consenting to participate. Participants were informed of participant confidentiality on the data collected. Demographic characteristics of the participants, including both age range and sex, were recorded as defining characteristics for the study population.

### Study Procedure

To avoid variability in the data collection, each participant was briefed and provided a demonstration of the correct application techniques for both TQ’s. Both the demonstration and briefing were consistent across all participants for the study. Following this, each participant was instructed to practice one application for both tourniquets on the TQ Aid. Participants were then informed that they would complete 3 trials for each tourniquet under time constraints. Furthermore, participants were briefed that they would alternate between both tourniquets throughout the duration of the data collection period.

Upon completion of the briefing period, participants began the series of applications starting with the CAT TQ. To increase standardization of applications across all participants, both tourniquets were set in the same position for each trial. Data points were then collected to follow predetermined time points in typical tourniquet application. Time began as soon as the participant picked the TQ off the table and began the process of applying the TQ Aid. The next data point collected was the time it took for participants to secure the TQ strap around the TQ Aid. Once secured, the next time point would be collected when the participant began twisting the windlass rod and pulling the ratchet for the CAT and Xforce TQ’s respectively. Lastly, a final time point was collected when the participant completed the application process, participant hands were removed from the TQ, and the TQ Aid indicated adequate pressure marked by a green LED light. Once the data collection was completed, participants were asked to record their age within categories of 10 years and sex. The consolidated standards of reporting trials (CONSORT) diagram for this study is presented in Figure 3.

**Figure 3.**
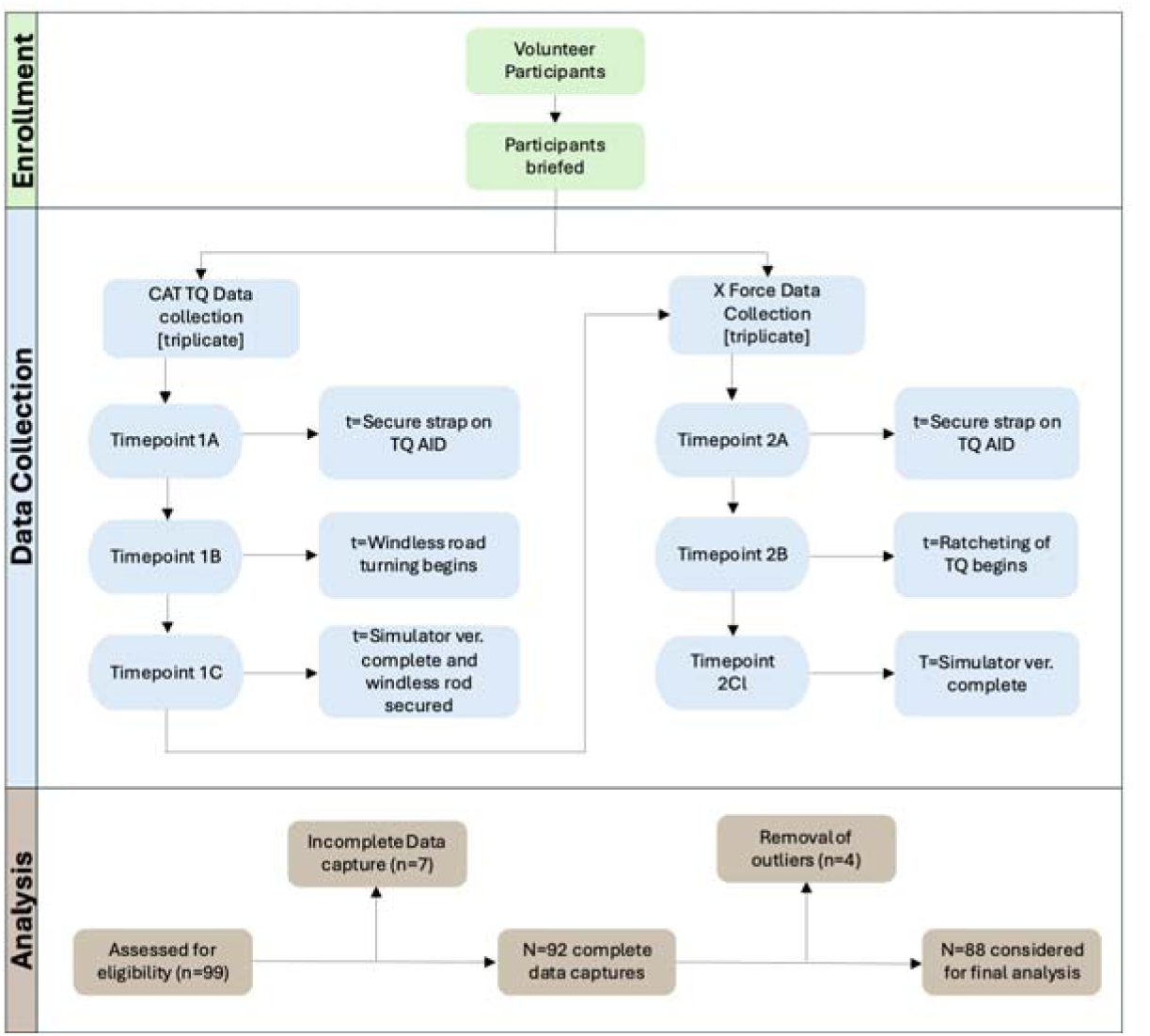
CONSORT Diagram of Study

**Figure 4.**
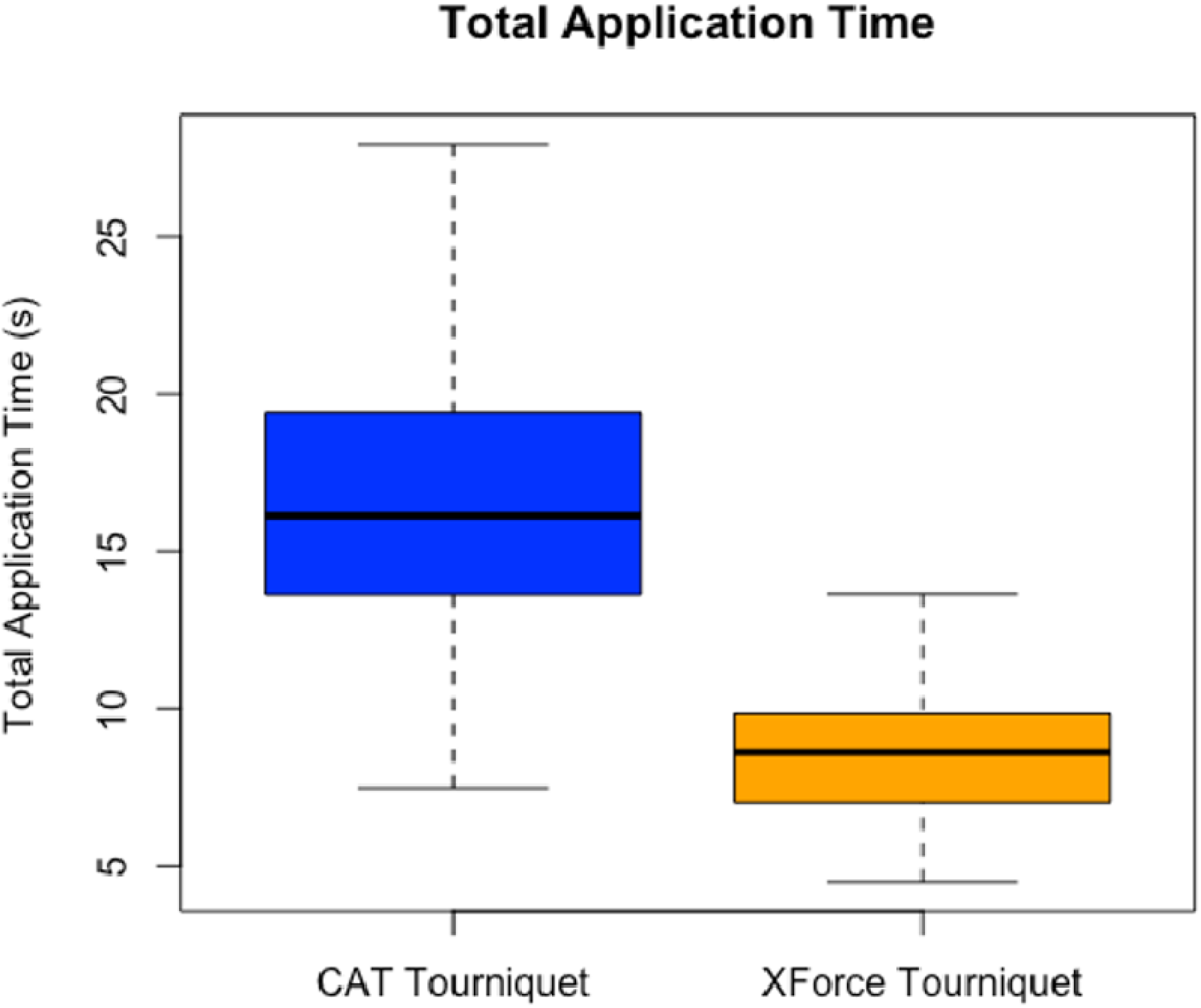
Comparison of mean total application time (seconds) between the CAT Tourniquet (16.53 ± 4.43) and XForce Tourniquet (8.67 ± 2.12), p < 0.001

**Figure 5.**
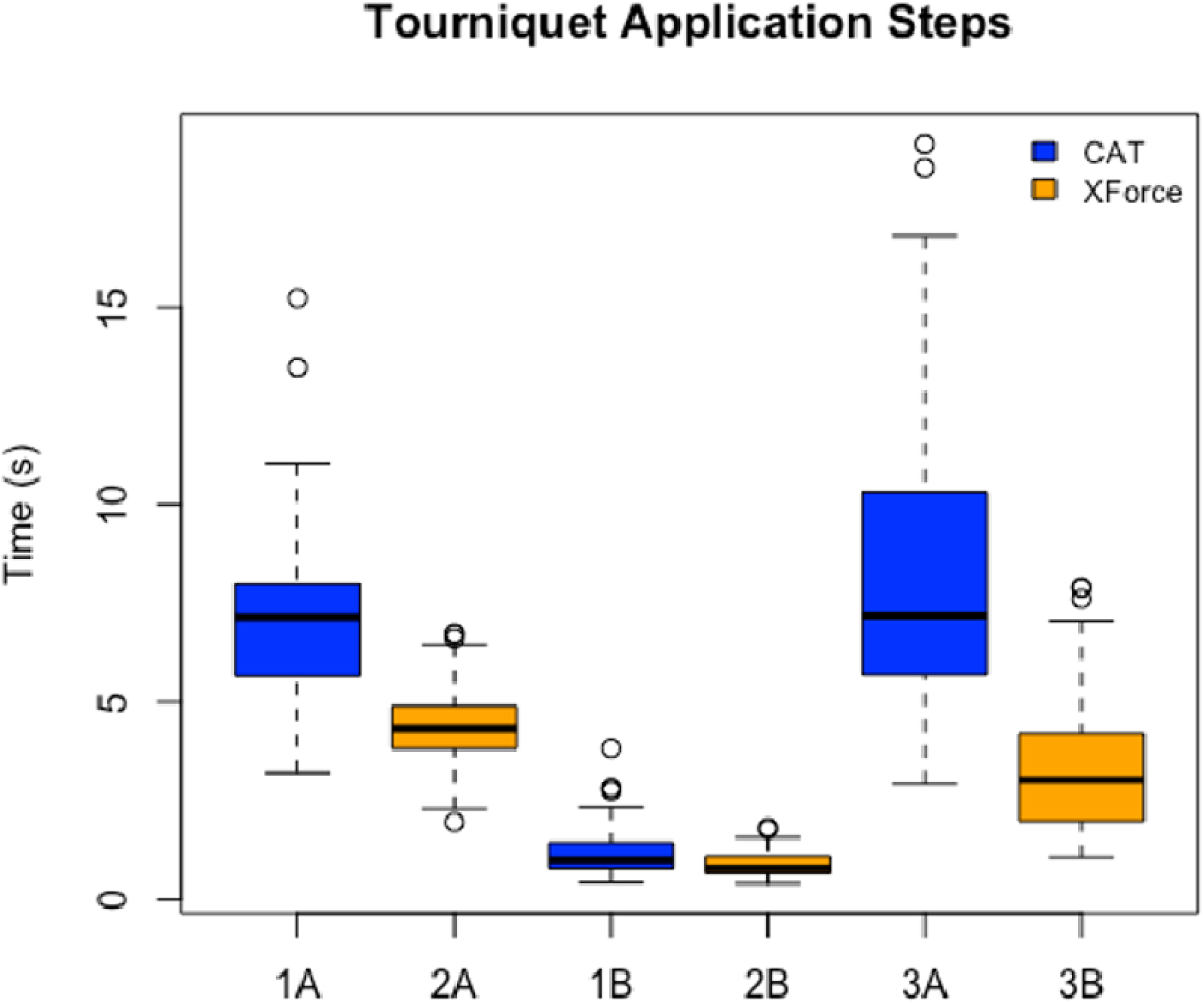
Comparison of mean time for tourniquet application steps between the CAT Tourniquet and XForce Tourniquet (reference Figure 3 for descriptions of timepoints 1A, 1B, 1C, 2A, 2B, 2C).

**Figure 6.**
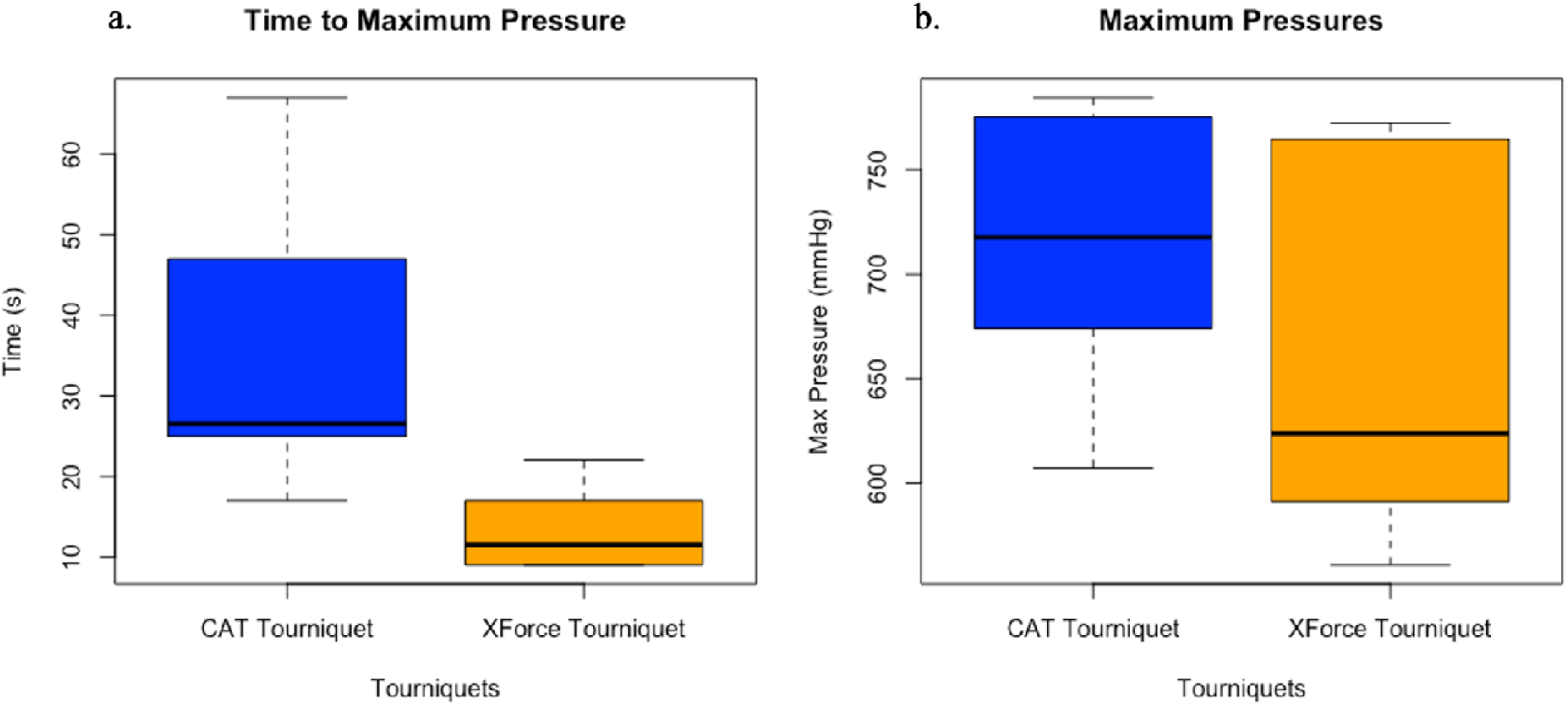
a) Comparison of mean time till maximum pressure (s) between the CAT Tourniquet (33.8 ± 15.89s) and XForce Tourniquet (12.9 ± 4.41s). b) Comparison of mean maximum pressure achieved between the CAT Tourniquet (712.87 ± 60.79 mmHg) and XForce Tourniquet (663.25 ± 87.70 mmHg), p= 0.1431.

## Results

### Validation of TQ Aid Pressure Sensor

To determine the validity of the TQ Aid pressure sensor, the Vernier Go Direct Gas Pressure Sensor was affixed to the TQ Aid and a total of n=20 applications were conducted. Both the CAT and XForce completed 10 trials each. The Vernier gas pressure sensor provided an average max pressure of 712.87 ± 60.79 and 663.25 ± 87.70 mmHg for the CAT and Xforce TQs respectively (Table 2). The average time to reach max pressure was 33.8 ± 15.89 and 12.9 ± 4.41 seconds for the CAT and Xforce TQs respectively. Evaluating results from a Mann-Whitney-Wilcoxon test (W= 70, p= 0.1431), at a 0.05 significance level, it can be concluded that there is no significant difference between maximum pressure achieved between CAT and XForce TQs.

**Table 1.**
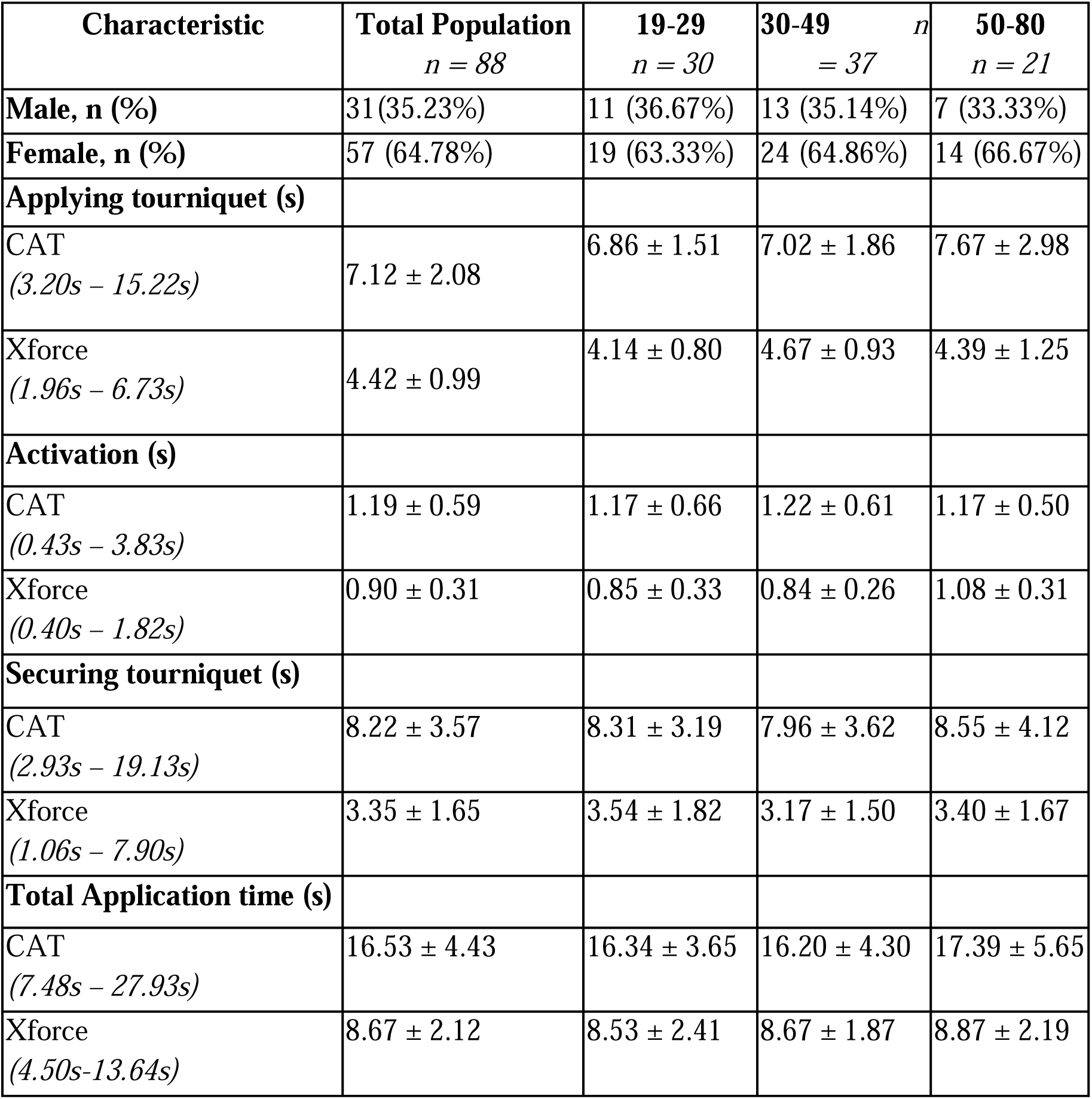
Baseline Characteristics of Age Specific Categories and Total Population

| Characteristic | Total Population<br>*n* = 88 | 19-29<br>*n* = 30 | 30-49<br>*n* = 37 | 50-80<br>*n* = 21 |
| --- | --- | --- | --- | --- |
| **Male, n (%)** | 31(35.23%) | 11 (36.67%) | 13 (35.14%) | 7 (33.33%) |
| **Female, n (%)** | 57 (64.78%) | 19 (63.33%) | 24 (64.86%) | 14 (66.67%) |
| **Applying tourniquet (s)** |  |  |  |  |
| CAT<br>(3.20s – 15.22s) | 7.12 ± 2.08 | 6.86 ± 1.51 | 7.02 ± 1.86 | 7.67 ± 2.98 |
| Xforce<br>(1.96s – 6.73s) | 4.42 ± 0.99 | 4.14 ± 0.80 | 4.67 ± 0.93 | 4.39 ± 1.25 |
| **Activation (s)** |  |  |  |  |
| CAT<br>(0.43s – 3.83s) | 1.19 ± 0.59 | 1.17 ± 0.66 | 1.22 ± 0.61 | 1.17 ± 0.50 |
| Xforce<br>(0.40s – 1.82s) | 0.90 ± 0.31 | 0.85 ± 0.33 | 0.84 ± 0.26 | 1.08 ± 0.31 |
| **Securing tourniquet (s)** |  |  |  |  |
| CAT<br>(2.93s – 19.13s) | 8.22 ± 3.57 | 8.31 ± 3.19 | 7.96 ± 3.62 | 8.55 ± 4.12 |
| Xforce<br>(1.06s – 7.90s) | 3.35 ± 1.65 | 3.54 ± 1.82 | 3.17 ± 1.50 | 3.40 ± 1.67 |
| **Total Application time (s)** |  |  |  |  |
| CAT<br>(7.48s – 27.93s) | 16.53 ± 4.43 | 16.34 ± 3.65 | 16.20 ± 4.30 | 17.39 ± 5.65 |
| Xforce<br>(4.50s-13.64s) | 8.67 ± 2.12 | 8.53 ± 2.41 | 8.67 ± 1.87 | 8.87 ± 2.19 |

**Table 2.** Pressure Sensor Readings for Tourniquets

| **Tourniquet** | **Avg Max**<br>**Pressure(mmHg)** | **Avg Time to Max**<br>**Pressure(s)** |
| --- | --- | --- |
| CAT (n=10) | 712.87 ± 60.79 | 33.8 ± 15.89 |
| Xforce (n=10) | 663.25 ± 87.70 | 12.9 ± 4.41 |

### Application of CAT and XForce TQs

Despite the recruitment of 99 participants for the study, only 92 completed all three trials for both the CAT and XForce TQs. Additionally, four outliers were identified using the interquartile range (IQR) method, resulting in a final sample size of 88 participants for statistical analysis. The final study population (n = 88) consisted of 31 males and 57 females. Participants were categorized into three age groups: 30 (9–29 years), 37 (30–49 years), and 21 (50–80 years). Regarding tourniquet application times, the mean total application time for the CAT and XForce tourniquets was 16.53 ± 4.43 s and 8.67 ± 2.12 s, respectively (Table 1). Mean total application time was calculated by averaging the three trials completed by each participant for each tourniquet.

Further, a paired t-test was conducted to determine whether there was a significant difference in mean total application time between the two tourniquets. The mean difference in total application time was 7.86 s, with the CAT TQ requiring significantly more time compared to the XForce TQ. Furthermore, at a 95% confidence level, the true mean difference in total application time was estimated to be between 7.03 s and 8.70 s. Overall, the XForce TQ demonstrated a significantly faster application time (8.67 ± 2.12 s) compared to the CAT TQ (16.53 ± 4.43 s, p < 0.001), reducing total application time by approximately 47%. These results strongly indicate a statistically significant difference in mean total application time between the CAT and XForce TQs.

To demonstrate consistency in application of both the CAT and XForce tourniquets between sexes, <u>2 two-sample t-tests</u> were conducted. Assumptions for a two-sample t-test were checked using a Shapiro-Wilk normality test & the Levene test for homogeneity of variance.

The first two-sample t-test (equal variance) was conducted to test the hypothesis that the mean total application time of the XForce tourniquet differs between sexes. The t-statistic was 2.65, with a p-value of 0.009. Thus, it can be concluded that there is a significant difference in mean total application time of the XForce tourniquet between males and females. The difference in mean total application time of the XForce tourniquet is 1.21 s, with a longer total application time in females (9.10 s) compared to males (7.88 s). With 95% confidence, the true mean difference in total application time could be as low as 0.30 s or as high as 2.12 s.

To determine if the difference in mean total application time of the CAT tourniquet between sexes is unique to the XForce tourniquet, another two-sample t-test (equal variance) was used to test the hypothesis that the mean total application time of the CAT tourniquet differs between sexes. The t-statistic was 2.29, with a p-value of 0.025. Thus, it can be concluded that there is a significant difference in mean total application time of the CAT tourniquet between males and females. The difference in mean total application time of the CAT tourniquet is 2.20 s, with a longer total application time in females (17.31 s) compared to males (15.11 s). With 95% confidence, the true mean difference in total application time could be as low as 0.28 s or as high as 4.12 s.

A one-way analysis of variance (ANOVA) test was used to determine if there is a significant difference between mean total application time of the XForce tourniquet between the three age groups (9-29 years, 30-49 years, 50-80 years). Assumptions for a one-way analysis of variance were checked using a Shapiro-Wilk normality test & the Levene test for homogeneity of variance. The F-value was not significant (F= 0.161, p= 0.852). Therefore, it can be concluded that there is no significant difference in mean total application time of the XForce tourniquet between the three age groups.

## Discussion

Tourniquets serve as a critical tool employed in the stoppage of hemorrhage for emergency medical services. Their crucial applications within emergency settings has been well-characterized in the military sphere and has recently gained attention in the civilian sector. As a result, research has focused on improving the efficacy of both application of TQs and the public’s general knowledge surrounding employing these devices in emergency situations. This study demonstrated that the newly developed XForce TQ offers significantly faster application times compared to the standard-of-care CAT TQ suggesting that its novel design features may enhance usability and efficiency in high-stress environments. Rapid hemorrhage control is critical in preventing exsanguination and requires users to be comfortable, efficient, and knowledgeable of TQ application. The findings of this study support that the XForce TQ provides additional comfort and efficiency when compared to the CAT TQ in a sample population of 99 healthcare volunteers.

The results of this study align with existing literature highlighting the challenges associated with the traditional windlass-based TQ design of the CAT TQ. Prior research has identified that even trained responders experience difficulty in securing and tightening windlass TQs due to their multi-step application process.^12,13^ This can contribute to prolonged application times and inconsistent pressure delivery. In contrast, the XForce TQ’s ratcheting lever mechanism and zip-tie strap system eliminate the need for a windlass, reducing the number of steps for proper application. Previous studies have reported mean CAT TQ application times to the upper limbs ranging from 44.53±13.47 to 51.96±16.96 seconds depending on different scenarios e.g., ordinary vs simulation scenarios.^14^ The XForce TQ, with an average application time of 8.67 seconds, represents a nearly 50% reduction in application time, suggesting a significant advantage in emergency scenarios. Reducing application time by nearly half may have profound clinical implications. Studies indicate that every minute of uncontrolled hemorrhage increases mortality risk^15,16^. The XForce TQ’s improved usability may therefore translate into improved patient outcomes in both civilian and military settings.

The ability to apply a TQ quickly and effectively serves to be particularly crucial in mass casualty incidents, military settings, and civilian trauma scenarios where time-to-treatment directly impacts survival outcomes. The design of the XForce TTQ addresses limitations of conventional TQ’s which serves to be particularly useful for untrained or infrequently trained users, a demographic that constitutes a significant portion of first responders in civilian emergencies. Through reducing the number of steps required for proper application, the XForce TQ has demonstrated potential in enhancing the effectiveness of public hemorrhage control initiatives such as the “Stop the Bleed” campaign. Additionally, the XForce TQ is engineered for efficiency in both application and manufacturing. The use of plastic injection molding allows for high-volume production with consistent quality. This seeks to address supply chain challenges that have historically affected the availability of emergency medical devices. The production process also provides scalability which is particularly relevant for disaster preparedness and military logistics.

Despite the promising findings portrayed in this study, several limitations must be considered. First, the study was conducted in a controlled, simulated environment. This environment does not replicate the complexity of real-world trauma scenarios. Such scenarios contain factors that affect TQ applications like patient movement, clothing barriers, and high-stress conditions which may influence application times. Future studies should develop methods to evaluate the XForce TQ in realistic pre-hospital and battlefield conditions to further validate its efficacy. Additionally, while this study assessed application speed, it did not directly measure applied pressure or occlusion success rates. While the TQ Aid simulator limb did provide a standardized pressure threshold (500 mmHg) for successful application, further research should explore whether the XForce TQ consistently achieves clinically advised occlusion pressures across diverse user populations. Future research should focus on testing the XForce TQ in real-world trauma settings rather than a simulated environment. Additionally, studies incorporating non-healthcare providers and lay responders will help evaluate the device’s usability across a broader population.

Despite the limitations of this study, the investigation conducted has provided evidence for the XForce TQs additional efficiency in comparison to the CAT TQ. The XForce, with faster and more consistent application times across age and sex, represents the foundational piece within a broader initiative to develop a next-generation intelligent tourniquet system. Future interactions will aim to integrate GPS tracking, automated emergency alerts, and telemedicine capabilities to enhance situational awareness and improve emergency response coordination. These features collectively aim to increase user comfort and confidence with TQ applications in emergency situations and more broadly aim to improve patient outcomes.

## Conclusion

This comparative study, conducted with human participants using a simulated limb approach, provides evidence that the XForce TQ significantly reduces application time compared to the standard-of-care CAT TQ. The XForce accomplishes this reduction in application time by employing a ratchet system which eliminates the traditional windlass mechanism of the CAT. The XForce also features an intuitive design, similar to that of zip-tie, which streamlines the application process thereby reducing the risk of user error and increasing the likelihood of effective induction of arterial occlusion during hemorrhagic crisis. While further research is needed to validate its performance in real-world trauma settings, this study provides compelling evidence that the XForce Tourniquet significantly reduces application time compared to the widely used CAT TQ, with implications for improved hemorrhage control in emergency settings. Thus, these findings lay the foundation for developing a next-generation intelligent tourniquet system incorporating smart features such as automated emergency alerts and telemedicine integration.

## Data Availability

All data produced in the present study are available upon reasonable request to the authors

## Abbreviations

TQ: Tourniquet
CAT: Combat Application Tourniquet
TCCC: Tactical Combat Casualty Care
ACS: American College of Surgeons

## Acknowledgements

This research was supported by grant funding from the New Jersey Commission on Science, Innovation, and Technology (CSIT) as part of its initiative to support New Jersey startups i.e., in this case Novara Solutions LLC. The Center for innovation at Rutgers Robert Wood Johnson Medical School and Robert Wood Johnson University Hospital provided location & resources for data collection & analysis.

## Authors Contributions

The study was conceptualized and designed by NY. AA, EP, and NY conducted the study, with AA and EP overseeing the entire data collection process. Data analysis was performed by AB & NY. AA & AB authored the initial manuscript. All authors contributed to the review process and have approved the final manuscript for submission.

## Declarations

### Ethics approval and consent to participate

This study was determined to be non-human research and, therefore, did not require institutional review board (IRB) approval. The study was conducted in accordance with ethical guidelines that prioritized confidentiality and integrity of the data.

### Competing interests

NY serves as an advisor, consultant, or board member for Turnkey Learning, LLC; Turnkey Learning (P) Ltd; Research Spark Hub; and Magnetic 3D. Additionally, Dr. Yanamala holds patents with Rutgers (US202163152686P; WO2022182603A1; US202163211829P; WO2022266288A1; US202163212228P; and WO2022266291A1) and with West Virginia University (invention numbers 2021-20 and 2021-047). All other authors (AA, EP and AB) have no relevant disclosures related to the contents of this paper.

## Notes

### Author Declarations

Ethics approval and consent to participate This study was determined to be non-human research and, therefore, did not require institutional review board (IRB) approval. The study was conducted in accordance with ethical guidelines that prioritized confidentiality and integrity of the data.

